# Tocilizumab in hospitalized patients with COVID-19: Clinical outcomes, inflammatory marker kinetics, safety, and a review of the literature

**DOI:** 10.1101/2020.08.05.20169060

**Authors:** Joshua A. Hill, Manoj P. Menon, Shireesha Dhanireddy, Mark M. Wurfel, Margaret Green, Rupali Jain, Jeannie D. Chan, Joanne Huang, Danika Bethune, Cameron Turtle, Christine Johnston, Hu Xie, Wendy M. Leisenring, H. Nina Kim, Guang-Shing Cheng

**Author notes:** Corresponding author. **Corresponding Author:** Joshua A. Hill, MD, Assistant Professor, Fred Hutchinson Cancer Research Center and University of Washington, 1100 Fairview Ave N, Mail Stop E-400, Seattle, WA 98109, Tele.

## Abstract

**Background:** Coronavirus disease 2019 (COVID-19) due to infection with SARS-CoV-2 causes substantial morbidity. Tocilizumab, an interleukin-6 receptor antagonist, might improve outcomes by mitigating inflammation.

**Methods:** We conducted a retrospective study of patients admitted to the University of Washington Hospital system with COVID-19 and requiring supplemental oxygen. Outcomes included clinical improvement, defined as a two-point reduction in severity on a 6-point ordinal scale or discharge, and mortality within 28 days. We used Cox proportional-hazards models with propensity score inverse probability weighting to compare outcomes in patients who did and did not receive tocilizumab.

**Results:** We evaluated 43 patients who received tocilizumab and 45 who did not. Patients receiving tocilizumab were younger with fewer comorbidities but higher baseline oxygen requirements. Tocilizumab treatment was associated with reduced CRP, fibrinogen, and temperature, but there were no meaningful differences in Cox models of time to clinical improvement (adjusted hazard ratio [aHR], 0.92; 95% CI, 0.38-2.22) or mortality (aHR, 0.57; 95% CI, 0.21-1.52). A numerically higher proportion of tocilizumab-treated patients had subsequent infections, transaminitis, and cytopenias.

**Conclusions:** Tocilizumab did not improve outcomes in hospitalized patients with COVID-19. However, this study was not powered to detect small differences, and there remains the possibility for a survival benefit.

## INTRODUCTION

Severe acute respiratory syndrome coronavirus 2 (SARS-CoV-2) is a novel human pathogen responsible for the largest global challenge to public health and humanity in over a century. Due to the high pathogenicity of this novel virus and a lack of pre-existing immunity, millions of individuals have been infected worldwide.(1) Among those infected, approximately 14% develop progressive pulmonary disease (coronavirus disease 2019 [COVID-19]) that results in critical illness in 5% of individuals.(2) Given high mortality rates in older patients and those with underlying comorbidities, there is an urgent need to identify effective therapies.

A profound inflammatory response to viral infection appears to be responsible for severe disease manifestations in patients with COVID-19. High serum levels of C-reactive protein (CRP), ferritin, and interleukin 6 (IL-6) are observed in patients requiring oxygen support and who progress to respiratory failure, cardiovascular collapse, and death and are consistent with a hyperinflammatory cytokine storm.(3–5) These laboratory and clinical findings are similar to the cytokine release syndrome (CRS) that may develop in cancer patients receiving chimeric antigen receptor immunotherapy (CAR-T cell therapy),(6) which has overlapping characteristics with macrophage activation syndrome (MAS).(7) Cytokine storm, and the resulting immunopathology, has also been identified as a cause of morbidity associated with related coronaviruses (SARS-CoV-1, Middle East Respiratory Syndrome-CoV) and other respiratory viruses.(8–11)

Treatment with corticosteroids, which suppress immune responses nonspecifically, has not been shown to improve outcomes with other severe respiratory viral infections or acute respiratory distress syndrome (ARDS) in the general population.(12) However, recent clinical trial data demonstrate a mortality benefit of low dose dexamethasone for COVID-19 in hospitalized patients requiring supplemental oxygen or mechanical ventilation,(13) supporting the notion that hyperinflammation contributes to disease pathogenesis among patients with severe COVID-19 disease. The increasing use of cytokine-specific immunomodulatory agents in other conditions has led to intense interest in the use of these agents for COVID-19. The rationale for use of immunomodulatory therapies in patients with COVID-19 is extrapolated from the experience in CAR-T cell therapy recipients, in whom IL-6 receptor blockade with tocilizumab (Actemra) can rapidly attenuate symptoms of CRS.(6) In this context, tocilizumab does not appear to independently increase risk for infection,(14, 15) although infectious complications are reported with chronic use in rheumatoid arthritis,(16) particularly with concurrent immunosuppressants. Cytokine blockade for hyperinflammatory syndromes is supported by additional lines of evidence, including the finding that treatment with recombinant IL-1 receptor antagonist, anakinra (Kineret), may improve outcomes in patients with MAS-like features in the setting of sepsis.(17)

Since the beginning of the pandemic, multiple observational reports have suggested that tocilizumab improves respiratory, radiological, and inflammatory parameters associated with COVID-19, although there are conflicting results with regard to clinical outcomes.(18–24) Observational studies using either another anti-IL-6 receptor antagonist, sarilumab (Kevzara), or anakinra have shown similar findings.(25–28) However, there are no published prospective randomized controlled trials that have proven efficacy for these anti-cytokine therapies for COVID-19. Furthermore, the safety of immunomodulatory therapy in the context of an active infection is not well understood. IL-6 is an essential aspect of immune control for many viral infections, but overproduction of IL-6 may be detrimental to viral clearance and survival.(10, 29) Thus, there is a critical need to understand the efficacy and safety of tocilizumab in patients with COVID-19. The aims of this observational study were to comprehensively describe the clinical course and outcomes of hospitalized patients with COVID-19 who were treated with tocilizumab and compare to patients from the same time period who were not treated with tocilizumab, describe adverse effects, and to evaluate the kinetics of inflammatory markers after tocilizumab therapy. In addition, we summarize the existing peer-reviewed literature regarding tocilizumab and COVID-19.

## METHODS

### Patients

We used the University of Washington (UW) Enterprise Data Warehouse to retrospectively identify all individuals admitted to three UW Hospitals across the Seattle metropolitan region between March 19^th^ (the first date of tocilizumab administration) and April 24^th^, 2020 with SARS-CoV-2 detection in a respiratory sample. Patients were excluded if they did not require supplemental oxygen, died or were placed on comfort measures within two days of admission (in the no tocilizumab cohort only), or received an alternative anti-cytokine therapy as part of clinical care or a trial. This study was approved by the UW Institutional Review Board.

### Treatment, supportive care, and monitoring

Hospital system-wide guidelines pertaining to the use of COVID-19-specific therapies were formulated using the limited available data by an interdisciplinary committee. During the study period, this guidance recommended use of open label hydroxychloroquine or investigational remdesivir in patients requiring supplemental oxygen and not participating on a corresponding clinical trial. The guidelines recommended consideration of a single dose of tocilizumab 400 mg intravenously in patients who had persistent fever and either impending or current respiratory failure, hemodynamic instability requiring vasopressor or ionotropic support, or a serum IL-6 level >5 times the upper limit of normal (range, 0-6 pg/mL). Tocilizumab was contraindicated in patients with sepsis due to another infection, transaminases >5 times the upper limit of normal, severe neutropenia (neutrophil count <500 cells/mm^3^), or severe thrombocytopenia (platelet count <50,000 cells/mm^3^). Corticosteroids and other antivirals were not recommended at that time. Patients received prophylactic heparin or enoxaparin for prevention of deep vein thrombosis at the discretion of the treating physician. Laboratory testing for inflammatory markers (IL-6, C-reactive protein [CRP], ferritin, fibrinogen, lactate dehydrogenase [LDH], and D-dimer) were recommended at the time of admission, every three days, with clinical deterioration, and prior to discharge.

### Outcomes

The primary outcome was sustained (≥3 days) clinical improvement within 28 days after baseline. For patients treated with tocilizumab, baseline was considered to be at the time of tocilizumab administration. For patients not treated with tocilizumab, baseline was considered to be 2 days after hospitalization to align with the median number of days between hospitalization and tocilizumab administration in the tocilizumab cohort (median, 1.8; IQR, 0.8-3.8). Clinical improvement was defined as a two-point reduction in patients’ baseline status on a six-point ordinal scale (modified from the WHO guidelines), or live discharge from the hospital, whichever came first. The ordinal scale consisted of 1, discharged; 2, hospitalized but not requiring oxygen; 3, hospitalized and requiring any supplemental oxygen; 4, hospitalized and requiring high-flow oxygen devices or non-invasive ventilation; 5, hospitalized and requiring invasive ventilation; and 6, death.(30) We also analyzed mortality within 28 days.

We described additional clinical outcomes of sepsis (defined as requiring hemodynamic support with vasopressors, with resolution at the time of vasopressor discontinuation), thrombotic events, bacteremia, and microbiologically confirmed pneumonia with another pathogen prompting antimicrobial therapy. We also described the kinetics of inflammatory laboratory markers, temperature, and heart rate.

### Data collection and statistical considerations

We abstracted data from medical records and electronic databases from the time of index admission for COVID-19 through 28 days after baseline as defined above. We described the proportion of patients with clinical improvement (as defined above) overall and stratified by baseline severity categories. We estimated the cumulative incidence of clinical improvement treating death as a competing risk. To mitigate differences in baseline characteristics between those who did and did not receive tocilizumab, we modeled the probability of treatment using baseline characteristics and calculated weights for inverse probability of treatment weighting (IPTW) analyses to weight the distribution of covariates to that of the overall population. Weights were standardized using the marginal probability of treatment and truncated at the 98^th^ percentile (values >4) to reduce variability.(31) We carried out multivariable Cox proportional-hazards modelling to evaluate the association of receipt of tocilizumab with 1) clinical improvement and with 2) mortality using IPTW and unweighted models. Robust variances were utilized in IPTW models to account for inclusion of the weights.(31) We also computed the restricted mean survival time (RMST) difference for time to clinical improvement and adjusted these models using IPTW. This analysis provides an intuitively appealing estimate of the difference in the mean number of days to clinical improvement between treatment groups.(32) Variables with a p-value <0.2 in univariable analyses were candidates for inclusion in the multivariable model; tocilizumab was included in all models, irrespective of univariate results. Variables were retained in the multivariable model if their inclusion modified the tocilizumab hazard ratio by more than 10% or if they were significantly associated with the outcome. In Cox models and RMST calculations for time to clinical improvement, patients who died before day 28 without clinical improvement were assigned a censored follow-up time at day 28, essentially giving them the worst possible “infinite” outcome. Additionally, we described the kinetics of changes in vital signs and laboratory results, as well as the occurrence of other clinical events including thrombosis and infections. SAS version 9.4 (SAS Institute, Cary, NC) and RStudio were used for analyses.

### Literature review

A literature search was conducted on July 10, 2020 via PubMed using the following search terms: (tocilizumab OR IL-6 OR interleukin-6) AND (COVID OR coronavirus OR SARS-CoV-2). Abstracts were reviewed to identify studies of individuals with confirmed SARS-CoV-2 infection who were treated with tocilizumab. Only studies with a comparator arm (i.e. no receipt of tocilizumab) were included.

## RESULTS

### Patient and treatment characteristics

Between March 19^th^, 2020 and April 24^th^, 2020, 44 patients were treated with tocilizumab. During this time period, an additional 120 patients were hospitalized with COVID-19 but did not receive tocilizumab. After excluding patients who did not require supplemental oxygen, who died or were placed on comfort measures within two days of admission, or who were enrolled in a placebo-controlled trial of IL-6 receptor blockade with sarilumab, there were 43 patients in the tocilizumab cohort and 45 patients in the non-tocilizumab cohort (**Figure S1**). Demographic and clinical characteristics of each cohort are shown in **Table 1** and stratified by baseline severity in **Table S1**. Compared to patients who did not receive tocilizumab, patients treated with tocilizumab were more likely to be younger, of white Hispanic race and ethnicity, without chronic lung disease, full code status, and receiving invasive ventilation (42% versus 20%) or high-flow oxygen (47% versus 22%). The invasive ventilation groups were similar overall. Most patients in both groups were treated with hydroxychloroquine, a minority (30%) were treated with remdesivir or placebo as part of a double-blind randomized clinical trial,(33) and 1 patient received open-label compassionate use remdesivir. No patients received corticosteroids for the treatment of COVID-19. The majority of patients received a single 400 mg intravenous dose of tocilizumab (median, 4.7mg/kg; **Table S2**); 3 patients received a second dose within 24 hours after the first.

**Table 1.**
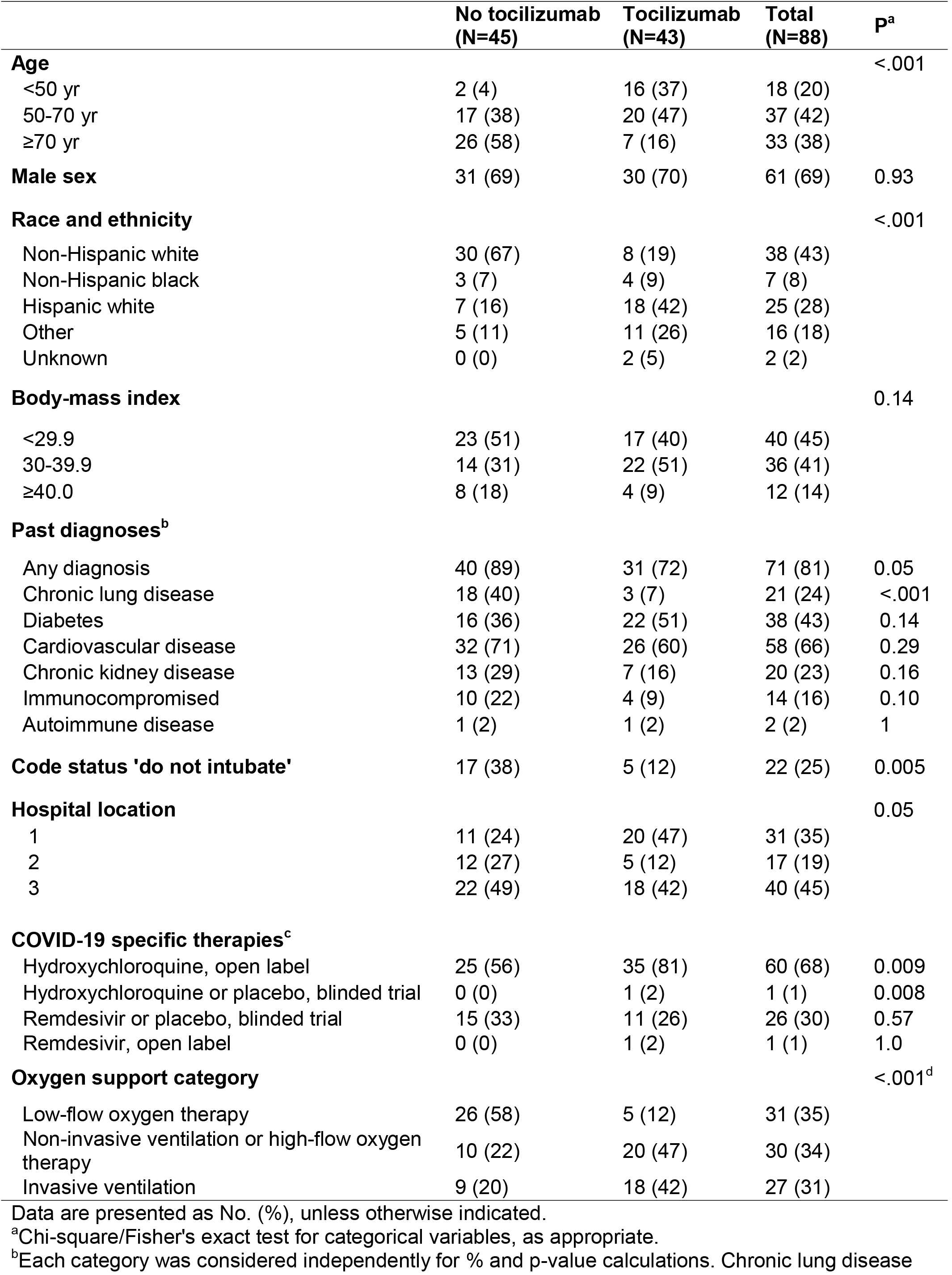
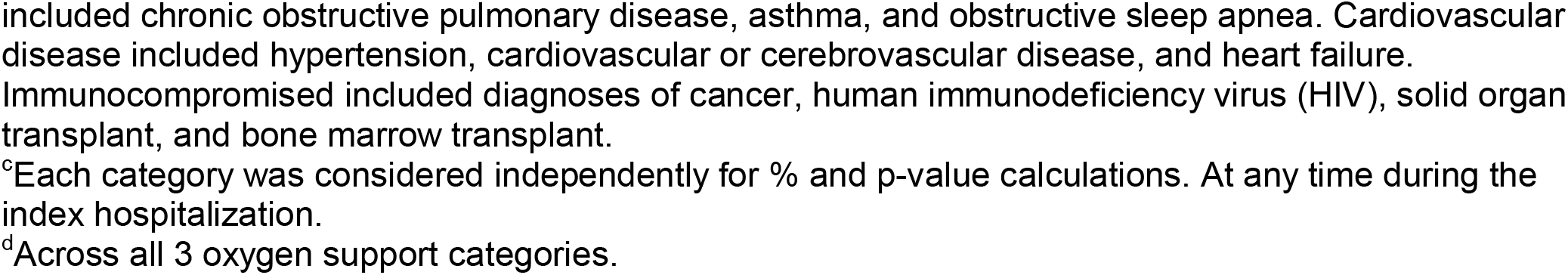
Baseline demographics and clinical characteristics.

### Clinical improvement

All patients were followed for up to 28 days after the baseline date or the time of death, whichever occurred first. In the tocilizumab cohort, 26 (60%) patients were discharged from the hospital, 9 (21%) died, and 8 (19%) remained hospitalized. In the no tocilizumab cohort, 27 (60%) patients were discharged from the hospital, 15 (33%) died, and 3 (7%) remained hospitalized. Among patients not requiring invasive ventilation at baseline and who did not have a ‘do not intubate’ code status, 5 of 15 (33%) and 0 of 3 (0%) patients progressed to invasive ventilation in the tocilizumab and no tocilizumab cohorts, respectively. The proportion of patients in each baseline severity category who met the clinical improvement endpoint are depicted in **Figure 1**, and longitudinal patient outcomes are depicted in **Figure S2**.

**Figure 1.**
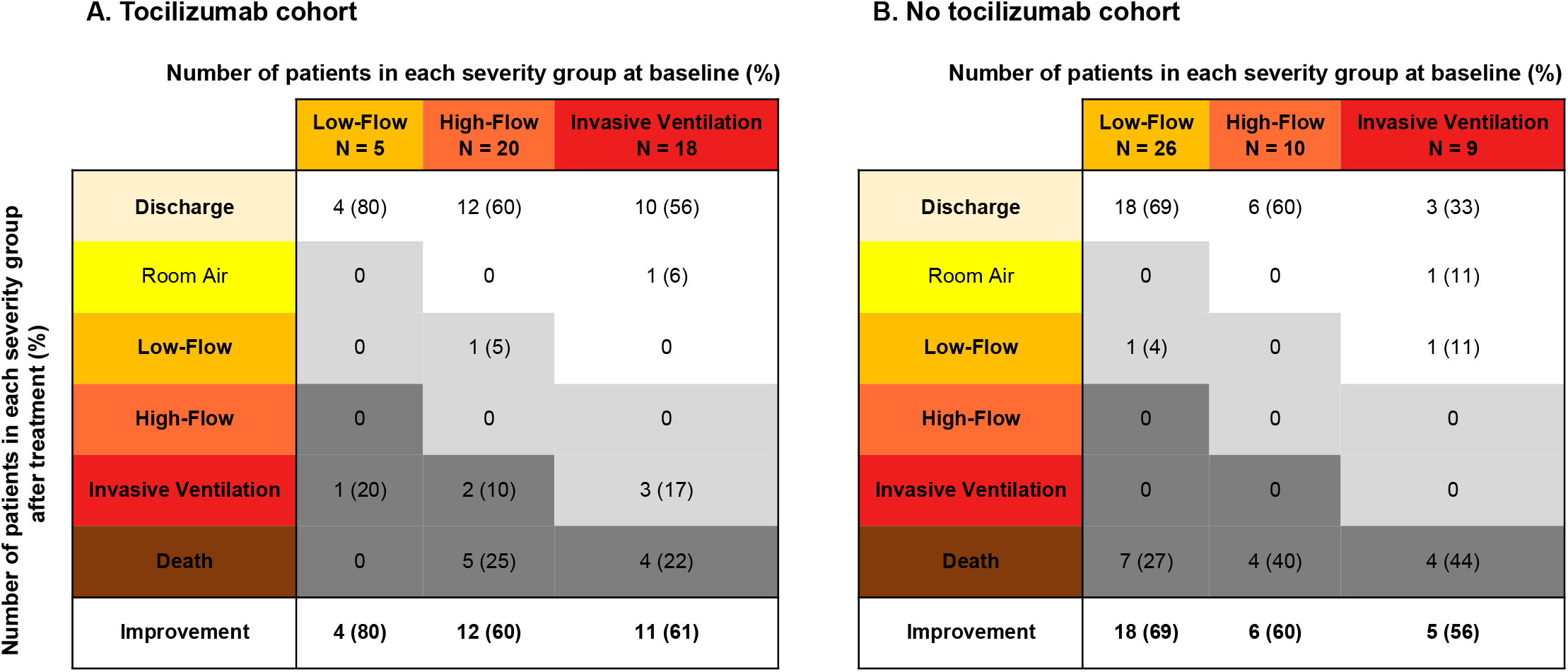
The proportion of patients in each baseline severity category who had clinical improvement by 28 days among patients in the tocilizumab and no tocilizumab groups. White boxes indicate clinical improvement for the indicated baseline severity category, light gray indicate neither improvement nor worsening, and dark grey indicate worsening. Percentages are column percentages based on the baseline severity category.

There were no differences in the cumulative incidence of clinical improvement among patients who did and did not receive tocilizumab overall or stratified by baseline severity categories (**Figure S3**). Among patients who improved, the median times to clinical improvement in the tocilizumab versus no tocilizumab cohorts, respectively, were 8 versus 6 days in the low-flow subgroup, 11 versus 9 days in the high-flow subgroup, and 16 versus 13 days in the invasive ventilation subgroup (**Table S3**).

To mitigate differences in baseline characteristics between tocilizumab treated and non-tocilizumab treated patients, we generated standardized IPTW weights from propensity scores (**Figure S4 A-B**). After truncating the two highest values, the IPT weighted population showed improvement in the balance of baseline characteristics between patients who did and did not receive tocilizumab (**Figures S4 C-D; Table S4**). In an adjusted Cox model using the truncated IPTW weights, receipt of tocilizumab was not associated with a difference in clinical improvement (adjusted hazard ratio [aHR], 0.92; 95% CI, 0.38-2.22; **Table 2**). Exploratory models within baseline oxygen categories had similar findings but were limited by small numbers within stratum (data not shown). There was also no evidence for a significant difference in the time to clinical improvement in an IPTW adjusted RMST analysis, which demonstrated that patients receiving tocilizumab improved an average of 1.08 days faster (RMST -1.08, 95% CI, -5.59 to 8.63 days, p=0.61). A cumulative incidence plot of time to clinical improvement using the IPT weighted population is shown in **Figure 2**.

**Table 2.** Multivariable Cox models of time to clinical improvement or death within 28 days using IPTW weights (truncated to account for outliers) to weight the distribution of covariates to that of the overall population.

| Variable | Adjusted hazard ratio<sup>a</sup> | P-value | 95% confidence interval |
| --- | --- | --- | --- |
| **Clinical improvement** |  |  |  |
| **Tocilizumab** |  |  |  |
| No | 1.00 |  |  |
| Yes | 0.92 | 0.85 | 0.38-2.22 |
| **Age group** |  |  |  |
| <50 yr | 1.00 |  |  |
| 50-70 yr | 1.81 | 0.18 | 0.76-4.27 |
| ≥70 yr | 1.35 | 0.67 | 0.33-5.44 |
| **Sex** |  |  |  |
| Male | 1.00 |  |  |
| Female | 0.95 | 0.92 | 0.37-2.44 |
| **Race and ethnicity** |  |  |  |
| Not non-Hispanic White | 1.00 |  |  |
| Non-Hispanic White | 0.58 | 0.29 | 0.21-1.59 |
| **Cardiovascular disease** |  |  |  |
| No | 1.00 |  |  |
| Yes | 0.46 | 0.08 | 0.19-1.11 |
| **Hospital** |  |  |  |
| 1 or 2 | 1.00 |  |  |
| 3 | 3.35 | 0.002 | 1.57-7.15 |
| **Code status 'do not intubate'** |  |  |  |
| No | 1.00 |  |  |
| Yes | 0.11 | <0.001 | 0.04-0.29 |
| **Oxygen support category** |  |  |  |
| Low-flow oxygen therapy | 1.00 |  |  |
| Non-invasive ventilation or high-flow oxygen therapy | 0.51 | 0.25 | 0.16-1.59 |
| Invasive ventilation | 0.15 | 0.002 | 0.05-0.49 |
| **Mortality** |  |  |  |
| **Tocilizumab** |  |  |  |
| No | 1.00 |  |  |
| Yes | 0.57 | 0.26 | 0.21-1.52 |
| **Age group** |  |  |  |
| <70 yr | 1.00 |  |  |
| ≥70 yr | 3.99 | 0.006 | 1.48-10.77 |
| **Race and ethnicity** |  |  |  |
| Not non-Hispanic White | 1.00 |  |  |
| Non-Hispanic White | 0.38 | 0.08 | 0.13-1.12 |
| **Hospital** |  |  |  |
| 1 or 3 | 1.00 |  |  |
| 2 | 2.44 | 0.11 | 0.82-7.29 |
| **Code status 'do not intubate'** |  |  |  |
| No | 1.00 |  |  |
| Yes | 10.61 | 0.003 | 2.18-51.71 |
| **Oxygen support category** |  |  |  |
| Low-flow oxygen therapy | 1.00 |  |  |
| Non-invasive ventilation or high-flow oxygen therapy | 2.65 | 0.09 | 0.87-8.0 |
| Invasive ventilation | 12.08 | 0.001 | 2.71-53.81 |

<sup>a</sup>A hazard ratio (HR) <1 indicates a lower likelihood of clinical improvement or mortality, as relevant to the model.

**Figure 2.**
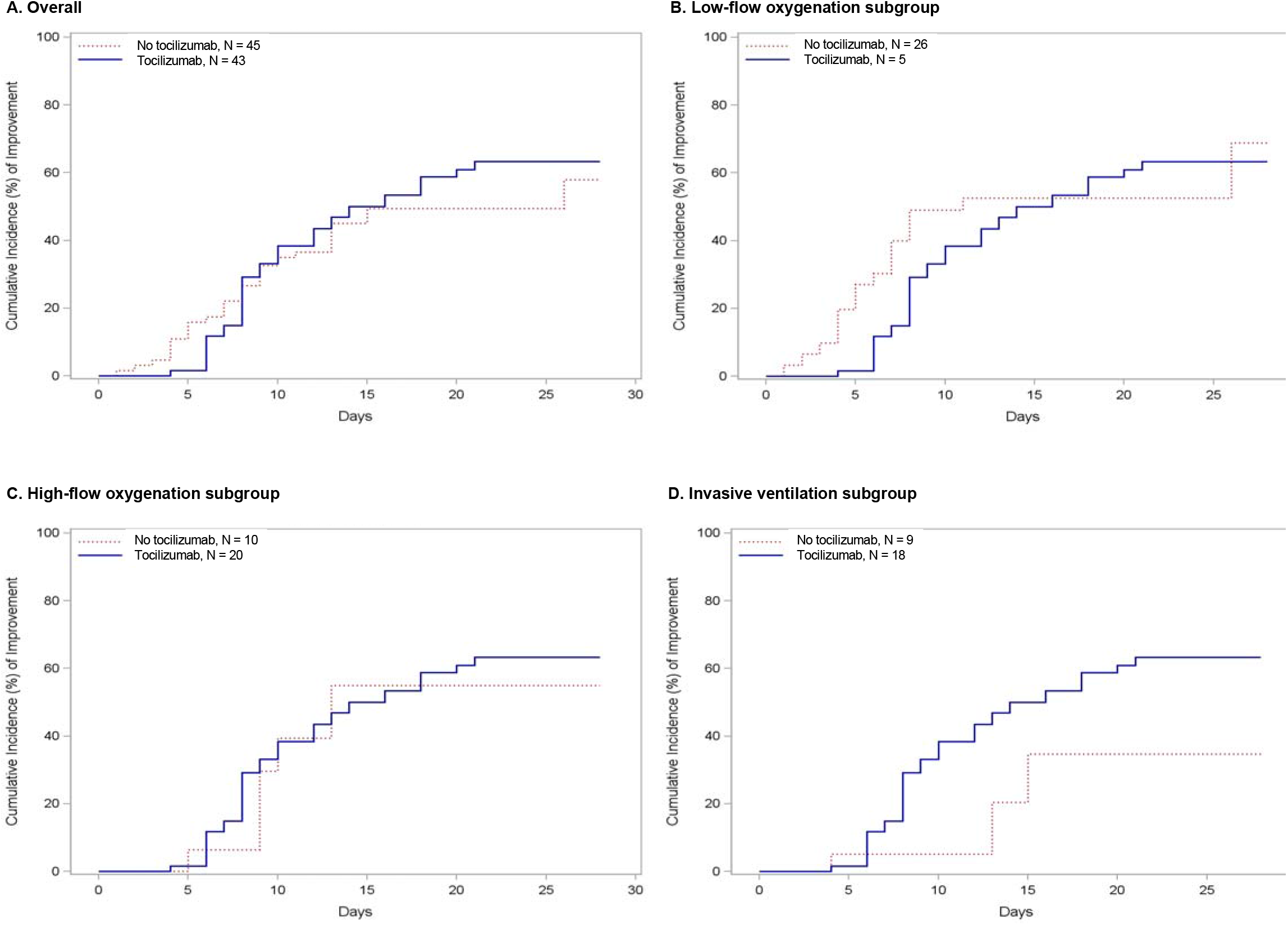
Cumulative incidence of sustained (≥3 days) clinical improvement within 28 days among patients who did and did not receive tocilizumab using the inverse probability of treatment (IPT) weighted population. Death was treated as a competing risk event.

### Mortality

An adjusted Cox model using the truncated IPTW weights demonstrated a lower observed risk of mortality within 28 days among patients treated with tocilizumab compared to those not treated with tocilizumab (aHR, 0.57; 95% CI, 0.21-1.52; **Table 2**). However, this estimate was highly variable and was not significant; as such, we cannot infer any difference in mortality between the treatment groups. A Kaplan-Meier survival curve using the IPT weighted population is shown in **Figure S5**.

### Other clinical outcomes

Sepsis within 1 day of tocilizumab or baseline date in the non-tocilizumab group occurred in 9 and 7 patients, respectively. Of the 9 tocilizumab patients, 7 survived (78%) with a median time to sepsis resolution of 3.7 days (IQR, 1.5-7.3); 2 patients died with ongoing sepsis 5 and 14 days after tocilizumab. Of the 7 non-tocilizumab treated patients, 3 survived (43%) with a median time to resolution of 13 days (IQR, 13-15); 4 patients died with ongoing sepsis at 1, 5, 12, and 14 days after baseline.

We also explored the potential benefit of tocilizumab for reducing thrombotic events. A numerically higher proportion of patients developed thrombotic events in the tocilizumab cohort (n=4, 9%) than in the non-tocilizumab cohort (n=2, 4%; **Table S5**). In addition, we evaluated potential adverse events associated with tocilizumab treatment. We found that a numerically higher proportion of patients in the tocilizumab cohort had bacteremia, secondary pneumonia, transaminitis, neutropenia, or thrombocytopenia within 14 days after baseline in the tocilizumab cohort (**Table 3**).

**Table 3.** Potential complications associated with tocilizumab.

|  | Tocilizumab cohort<br>(N = 43) | No tocilizumab cohort<br>(N = 45) |
| --- | --- | --- |
| Bacteremia<sup>a</sup> | 4 (9)<sup>b</sup> | 2 (4)<sup>c</sup> |
| Ventilator-associated pneumonia<sup>d</sup> | 9 (21)<sup>e</sup> | 5 (11)<sup>f</sup> |
| AST or ALT >5x baseline<sup>g</sup> | 4 (9) | 2 (4) |
| Absolute neutrophil count <1,000 cells/mm<sup>3h</sup> | 0 (0) | 0 (0) |
| Platelet count <100,000 cells/mm<sup>3i</sup> | 5 (12) | 2 (4) |

Data are presented as number of individuals (%) and represent new events that occurred within 14 days of the baseline date.
<sup>a</sup>Defined as positive blood cultures that were treated with antibiotics.
<sup>b</sup>Pathogens consisted of coagulase-negative Staphylococci (CONS), *Serratia marscecens*, and *Enterococcus faecalis*.
<sup>c</sup>Pathogens consisted of methicillin-sensitive *Staphylococcus aureus* (MSSA), *Granulicatella adiacen*, and CONS.
<sup>d</sup>Defined as microbiologic documentation in addition to antimicrobial therapy. All events occurred in patients who were mechanically ventilated in both cohorts.
<sup>e</sup>Pathogens consisted of MSSA, *Haemophilus influenzae*, *Escherichia coli*, *Serratia marscecens*, methicillin-resistant *Staphylococcus aureus* (MRSA), *Klebsiella pneumoniae*, and *Klebsiella variicola*.
<sup>f</sup>Pathogens consisted of *Haemophilus influenzae*, *Sterptococcus pneumoniae*, MSSA, *Enterobacter aerogenes*, and *Serratia marcescens*.
<sup>g</sup>7 patients did not have any results, and 11 patients did not have any results after baseline.
<sup>h</sup>1 patient did not have any results, and 7 patients did not have any results after baseline.
<sup>i</sup>5 patients had an absolute neutrophil count <1,000 cells/mm<sup>3</sup> at baseline; 3 patients did not have any results after baseline.

### Effect of tocilizumab on clinical and laboratory markers of inflammation

Laboratory values were similar at baseline for the tocilizumab and no tocilizumab cohorts (**Figure 3**). Among most patients who received tocilizumab, there was a rapid and sustained decrease in temperature (**Figure S6**), CRP, and fibrinogen. There was an initial increase followed by a decrease in IL-6 levels after administration of tocilizumab as expected given the mechanism of action of blocking the IL-6 receptor. Levels of LDH declined over days, and there was minimal change in ferritin and D-dimer levels. Heart rates rapidly declined but changes were not sustained over time (**Figure S6**). In contrast, levels of all biomarkers were relatively stable in the no tocilizumab cohort.

**Figure 3.**
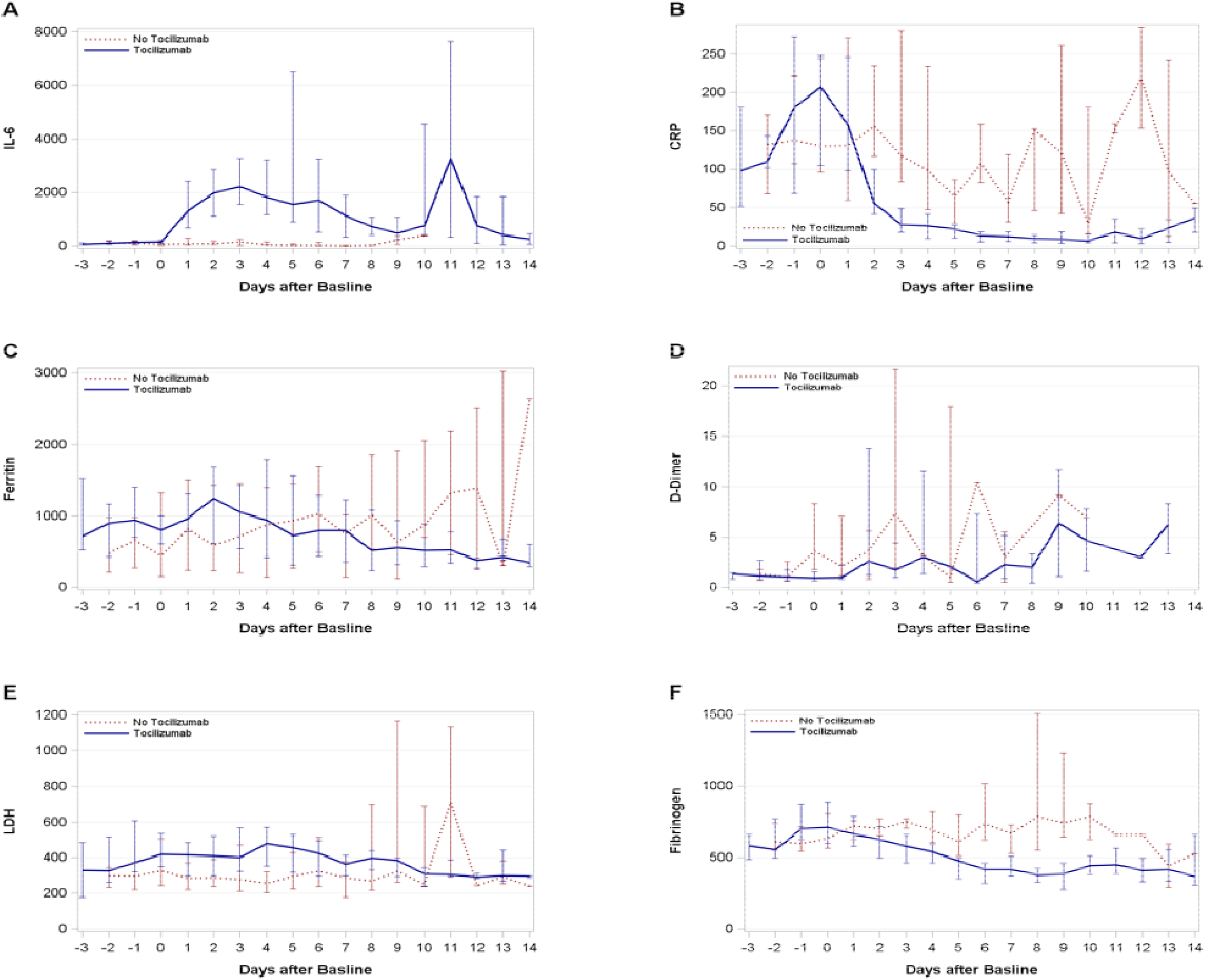
Kinetics of inflammatory laboratory markers. IL-6 indicates interleukin 6 (pg/mL); CRP, C-reactive protein (mg/L); LDH, lactate dehydrogenase (U/L). Units for other laboratory values are ng/mL for ferritin, mcg/mL for D-dimer, mg/dL for fibrinogen. The presented data indicate medians and the associated interquartile ranges. If multiple values were available for the same day, the maximum value was used. The number of patients contributing results to each panel are in **Table S6**. In Panel A, the late secondary increase in IL-6 levels was driven by 2 individuals who had progressive cardiopulmonary disease with concurrent bloodstream infections and ventilator-associated pneumonias at the time of secondary IL-6 increase.

## DISCUSSION

Morbidity and mortality remain unacceptably high in patients with COVID-19. The clinical utility of directed immunomodulatory agents to preempt or reverse the cytokine-driven inflammation responsible for much of this morbidity has biological plausibility,(34) but there are limited data to guide treatment strategies. Although tocilizumab resulted in normalization of multiple inflammatory biomarkers, we did not find clear evidence that tocilizumab improved clinical outcomes, although we were not powered to detect small effects, and we cannot rule out a survival benefit. There were numerically more infections, transaminitis, and cytopenias in individuals treated with tocilizumab.

Published observational studies have provided mixed results in terms of the associated benefits of tocilizumab treatment in hospitalized patients with COVID-19 (**Table 4**). Our literature review of studies with a comparator arm illustrate substantial heterogeneity in patients, their treatments, definitions of outcomes, and analytic approaches. Most studies, including ours, are limited by residual indication bias among those receiving tocilizumab. Rapidly evolving improvements in supportive care strategies may also confound findings of better outcomes with tocilizumab over time. In our study of patients with heterogeneous disease severity, we did not demonstrate an apparent clinical benefit of treatment with tocilizumab. Patients receiving tocilizumab in our study were younger and sicker on presentation, and thus more likely to receive aggressive care early in the pandemic. Our study was not confounded by the use of corticosteroids, unlike most other observational studies. This is relevant given emerging data of the benefit of corticosteroids in patients with severe disease.(35). The possibility still remains that there is a clinical scenario in which tocilizumab is beneficial, but this will be best demonstrated through randomized clinical trials.

**Table 4.** Peer-reviewed retrospective studies with comparator groups of clinical outcomes in patients treated with tocilizumab for COVID-19.

|  | Invasive ventilation | Toci dose | Antivirals used in majority | Steroid use | Clinical improvement | Mortality | Secondary infections | Other reported adverse effects |
| --- | --- | --- | --- | --- | --- | --- | --- | --- |
| **Guaraldi (44)**<br>Toci: N = 179<br>No Toci: N=365 | Toci: 18%<br>No Toci: 16% | 8 mg/kg x 2 IV or 162 mg x 2 SC | HCQ, Azithro, and L/R or D/C | Toci: 30%<br>No Toci: 17% | Toci: 23% died or mechanically ventilated<br>No Toci: 37% died or mechanically ventilated by day 14 | Toci: 7%<br>No Toci: 20% | Toci: 13%<br>No Toci: 4% | Severe neutropenia (n=1), injection site reaction (n=1) |
| **Somers (24)**<br>Toci: N = 78<br>No Toci: N = 76 | Toci: 100%<br>No Toci: 100% | 8 mg/kg x 1 | None | Toci: 29%<br>No Toci: 20% | Toci: 45% reduction in hazard for death | Toci: 18%<br>No Toci: 36% (by day 28) | Toci: 54%<br>No Toci: 26% | None reported |
| **Rojas-Marte (45)**<br>Toci: N = 96<br>No Toci: N = 97 | Toci: 64%<br>No Toci: 62% | Not reported | HCQ, Azithro | Toci: 43%<br>No Toci: 33% | Length of hospital stay:<br>Toci 14.5 ± 8.8 days,<br>No Toci: 16.5 ± 10.8 days | Toci: 45%<br>No Toci: 57% | Toci: 13% bacteremia, 4% fungemia<br>No Toci: 24% bacteremia, 3% fungemia | None reported |
| **Capra (19)**<br>Toci: N = 62<br>No Toci: N = 23 | 0 | 400 mg x 1 IV or 324 mg SC | HCQ, L/R | No | Toci: 92%<sup>a</sup><br>No Toci: 42%<sup>a</sup> complete recovery | Toci: 8%<sup>a</sup><br>No Toci: 58%<sup>a</sup> | None | Not reported |
| **Campochiaro (46)**<br>Toci: N = 32<br>No Toci: N = 33 | 0 | 400 mg x 1, no second doses | HCQ, Azithro, L/R | No | Toci: 69%<br>No Toci: 61% improvement by 28 days | Toci: 15%<br>No Toci: 33% (by day 28) | Toci: 13%<br>No Toci: 12% | Transient increase in AST/ALT (Toci: 15%, No Toci: 18%); transient neutropenia (Toci: 16%, No Toci: 0%) |
| **Colaneri (47)**<br>Toci: N = 21<br>No Toci: N = 21 | Not reported | 8 mg/kg<br>x 2 | HCQ,<br>Azithro | All subjects | No difference in risk for ICU admission or death by 7 days | Toci: 24%<br>No Toci: 21% (by day 7) | None | None |
| **Klopfenstein (22)**<br>Toci: N = 20<br>No Toci: N = 25 | Not reported | Not reported | HCQ, L/R | Not reported | Toci: 25% died or admitted to ICU<br>No Toci: 72% died or admitted to ICU | Toci: 25%<br>No Toci: 48% | Not reported | Not reported |
| **Quartuccio (23)**<br>Toci: N = 42<br>No Toci: N = 69 | Toci: 62%<br>No Toci: 0 | 8 mg/kg<br>x 1 | HCQ, L/R or D/C | Toci: 38%<br>No Toci: 0 | Toci: 71%<br>No Toci: 100% clinical improvement | Toci: 10%<br>No Toci: 0% | Toci: 43%<br>No Toci: 0% | Not reported |
| **Price (48)**<br>Toci: N = 153<br>No Toci: N = 86 | Toci: 31%<br>No Toci: 6% | 8 mg/kg<br>x 1, max 800 mg | HCQ | 20% overall | Toci: 86%<br>No Toci: 87% 14-day survival | Toci: 14%<br>No Toci: 13% by day 14 | Toci: 3% (bacteremia) | Toci: 4% neutropenia |
| **Kewan (43)**<br>Toci: N = 28<br>No Toci: N = 23 | Toci: 75%<br>No Toci: 48% | 400 mg<br>x 1 | HCQ, Azithro | Toci: 71%<br>No Toci: 48% | Shorter time to clinical improvement | Toci: 14%<br>No Toci: 18% | Toci: 18%<br>No Toci: 22% | None |
| **Potere (49)**<br>Toci: N = 40<br>No Toci: N = 40 | Toci: 0%<br>No Toci: 0% | 324 mg<br>x 2 SC | HCQ | Toci: 65%<br>No Toci: 58% | Toci: 93%<br>No Toci: 73% improvement on ordinal scale by day 35 | Toci: 5%<br>No Toci: 28% by day 35 | Toci: 3%<br>No Toci: 8% | Toci: ALT rise (3%) |

N indicates number; HCQ, hydroxychloroquine; L/R, lopinavir/ritonavir; D/C, darunavir/cobicistat; azithro, azithromycin; IFN, interferon; toci, tocilizumab; SC, subcutaneous.
<sup>a</sup>Among those who met an endpoint of discharge from the hospital or death, which was met by 25 of patients in the tocilizumab group and 19 in the control group.
It is important to note that tocilizumab dose and administration timing, as well as definitions of clinical improvement, secondary infections, and adverse events varied by study.

We also tested the novel hypothesis that thrombotic events may be reduced in tocilizumab-treated patients. This could be an additional benefit of tocilizumab based on the high frequency of venous thrombosis in patients with COVID-19.(36) It is biologically plausible that tocilizumab could reduce risk for thrombosis given studies demonstrating reduced plasma levels of the procoagulant Factor XIII (fibrin-stabilizing factor), d-dimer levels, and other inflammatory markers in patients receiving tocilizumab.(37–40) Although we demonstrate a clear anti-inflammatory effect with rapid and marked reductions in serum levels of CRP and fibrinogen, a higher proportion of patients in the tocilizumab group in our study had thrombotic events, which may be due to the higher baseline disease severity in the tocilizumab group. It is possible that thromboembolic events were underdiagnosed overall during the study period.(36, 41) The low number of events and differences in characteristics between the patient subgroups limit conclusions, but the role of tocilizumab for reducing thrombosis may warrant additional study in randomized trials.

Due to the potential for increased infection risk with chronic use of tocilizumab in patients with rheumatoid arthritis,(16) we evaluated secondary infections. Although a numerically higher proportion of patients in the tocilizumab cohort had a microbiologically confirmed secondary infection, this was likely related to more patients requiring invasive ventilation in the tocilizumab group, resulting in more diagnoses of ventilator-associated pneumonias. Some studies report higher rates of infections in patients receiving tocilizumab (**Table 4**), although this may not compromise the potential benefits of tocilizumab.(24) We also report numerically higher rates of transaminitis and cytopenias in patients receiving tocilizumab. These comparisons were all limited by a small number of events and may reflect differences in the distribution of disease severity between the two groups. Our findings do not suggest obvious safety concerns related to tocilizumab use for COVID-19, consistent with the majority of studies.

Finally, we evaluated the effect of tocilizumab on clinical and laboratory markers of inflammation. Similar to other studies of tocilizumab in patients with cytokine release syndrome and COVID-19,(18, 42) we demonstrated a rapid and sustained effect on lowering body temperature, CRP, and fibrinogen. This may play a role in the higher rates of resolution of sepsis physiology that we describe in our cohort and as reported in another study.(43) The anti-inflammatory effects also suggest that the ~4 mg/kg dose used in our patients was sufficiently bioactive. Whether the kinetics of inflammatory biomarkers correlate with response to tocilizumab warrants further study.

The primary limitations of our study include those inherent to a retrospective, non-randomized design. Higher baseline severity status in the tocilizumab cohort, as well as other differences in patient characteristics, resulted in imbalances in the patient cohorts. To control for some of these differences, we compared outcomes within baseline severity categories and performed IPTW adjusted analyses. For most comparisons, we were limited to detecting relatively large differences. Strengths of this study include the relatively large cohort of patients requiring noninvasive and invasive oxygenation, along with careful adjustment for differences in patient characteristics and other confounders. We performed an RMST analysis for time to clinical improvement that is novel to studies of tocilizumab for COVID-19 and provide a tangible measure of treatment effect.(32) We performed comprehensive chart review and data abstraction to describe and analyze a variety of clinical and laboratory outcomes to evaluate the potential benefits and side effects of tocilizumab therapy, including novel comparisons of thrombotic events and resolution of sepsis physiology. Our data were not confounded by steroids, and most patients did not receive remdesivir.

In conclusion, there was no clear evidence of improved clinical outcomes among hospitalized patients with COVID-19 who received one dose of 400 mg of tocilizumab without concurrent steroids or effective antivirals. Tocilizumab was associated with a robust reduction in inflammatory biomarkers; further study in patients with sepsis physiology may be warranted. Randomized controlled trials using tocilizumab in the context of concurrent effective antiviral and steroid therapy will be particularly important to guide evolving treatment strategies.

## Data Availability

Data is available upon request

## Acknowledgements

We acknowledge the University of Washington Division of Allergy and Infectious Diseases Research Collaboratory, including Ayushi Gupta and Kristine Lan, for help with data abstraction and analysis. We acknowledge Rina Romano and Joyce Maalouf for help with chart review, as well as Aimee Forquera and Linda Lei for patient care coordination. We appreciate the assistance of Ashley Sherrid for the literature review.

## Author contributions

J.A.H., M.M., H.N.K., and GS.C designed the study and interpreted the data. J.A.H., M.M., S.D., M.G., R.J., J.C., J.H., and GS.C. contributed to data collection. J.A.H., H.N.K., H.X. and W.M.L. analyzed the data and created the figures. J.A.H. drafted the manuscript. All authors contributed to the writing and revision of the manuscript and approved the final version.

